# Modeling Hospital Energy and Economic Costs for COVID-19 Infection Control Interventions

**DOI:** 10.1101/2020.08.21.20178855

**Authors:** Marietta M. Squire, Megashnee Munsamy, Gary Lin, Arnesh Telukdarie, Takeru Igusa

## Abstract

The objective of this study was to assess the energy demand and economic cost of two hospital-based COVID-19 infection control interventions. The intervention control measures evaluated include use of negative pressure (NP) treatment rooms and xenon pulsed ultraviolet (XP-UV) infection control equipment. After projecting COVID-19 hospitalizations, a Hospital Energy Model and Infection De-escalation Models are applied to quantify increases in energy demand and reductions in secondary infections. The scope of the interventions consisted of implementing NP in 11, 22, and 44 rooms (at small, medium, and large hospitals) while the XP-UV equipment was used eight, nine, and ten hours a day, respectively. The annum kilowatt-hours (kWh) for NP (and costs were at $0.1015 per kWh) were 116,700 ($11,845), 332,530 ($33,752), 795,675 ($80,761) for small, medium, and large hospitals ($1,077, $1,534 $1,836 added annum energy cost per NP room). For XP-UV, the annum kilowatt-hours and costs were 438 ($45), 493 ($50), 548 ($56) for small, medium, and large hospitals. There are other initial costs associated with the purchase and installation of the equipment, with XP-UV having a higher initial cost. XP-UV had a greater reduction in secondary COVID-19 infections in large and medium hospitals. NP rooms had a greater reduction in secondary SARS-CoV-2 transmission in small hospitals. Early implementation of interventions can result in realized cost savings through reduced hospital-acquired infections.

## 1. Introduction

Hospitals are inherently energy intensive buildings. Hence, hospital operations require the assessment of energy efficiency when evaluating Coronavirus Disease 2019 (COVID-19) intervention measures, due to COVID-19 being caused by the Severe Acute Respiratory Disease Coronavirus-2 (SARS-CoV-2). Given environmental and sustainability concerns, studies have worked on assessing the energy demands of hospitals under normal circumstances [1]. On a global scale, energy demands have plummeted during the COVID-19 pandemic [2]. But for the healthcare sector, depending on the geographic area, energy demands may be higher than usual. Access to energy will be paramount for critical healthcare delivery and access, especially in developing countries [3].

In addition to analyzing energy consumption, the safety performance of hospitals is also of paramount importance. Hospital safety should be accounted for since many hospitals have a mission to maintain stringent protocols to keep patients and staff safe along with providing a functional environment for healthcare workers to perform their duties. Higher connectivity of countries and increased transportation within the last 50 years have exacerbated the incidences of infectious diseases and increased the burden on hospital systems. This has particularly been evident in the recent COVID-19 pandemic [4]. According to the World Health Organization, as of June 12, 2020, 215 countries and territories had confirmed SARS-CoV-2 infections [5].

Hospital preparation and response to outbreak and epidemic events are important for both patient safety and to protect healthcare workers. Given the transmissibility of SARS-CoV-2, supplements to existing infection control measures will incur excess energy on existing hospital usages, since various disease transmission mitigation precautions require augmentation to the existing hospital infrastructures. Furthermore, additional infrastructure costs to the hospital facility, such as improvements to ventilation, will scale-up to impact healthcare system-level costs as well as costs of care to the patient and payers.

Hospital facilities have unique requirements compared to other buildings given that patient outcomes (patient pain levels, lengths of stay, infection rates, patient recovery rates, patient mortality rates) as well as staff outcomes (such as overall workplace safety, infection control metrics, stress levels, and staff sense of well-being), are intricately connected to the built environment of the hospital itself [6-7]. Additionally, indoor air quality and HVAC parameters can impact patient outcomes [7]. Air flow and thermal control can directly relate to patient comfort and recovery and hospital patient satisfaction metrics [8]. Singh et al. (2017) states some of the energy challenges, among many, associated with hospitals include: 24-hour operations, varied space use, and the use of energy intensive equipment [9]. Inadequate lighting and ‘thermal discomfort’ can influence and make medical diagnosis more error prone [10]. Balancing the priorities of a ‘sustainable design’ and creating an environment that is conducive to patient recovery and staff wellness and safety is difficult to achieve [6,8]. Hence, we developed a methodology that investigated hospital energy demands and secondary transmission of the SARS-CoV-2 pathogen specifically in the COVID-19 pandemic context.

This study provides a framework to evaluate increases in energy demand, cost, reduction in secondary infections, and potential cost savings of implementing infection control measures (which require additional energy use) in a hospital setting. We apply optimization techniques to assess specifically how budgetary funds minimize secondary infections with the least financial impact. The aims are to: (1) Quantify both normal and COVID-19 energy scenario demands and costs, (2) quantify the efficacy of the COVID-19 infection control interventions, and (3) optimize cost allocations across the interventions. Based on an annual projected window of hospitalizations in the state of Maryland, three hospitals with varying sizes were evaluated.

## 2. Review of Energy Use and Interventions

### 2.1 Hospital Energy Usage

Lombard et al. (2008) reported that hospitals account for 9% (United States), 11% (Spain), and 6% (United Kingdom) of commercial energy use in these respective countries [11]. Hospitals in the United States have an average energy use of 786 kWh per square meter per year, second to restaurants, which have an average energy usage of 814 kWh per square meter per year [11]. According to the 2012 US Commercial Building Energy Consumption Survey (CBECS) a typical hospital has more floor area than an average restaurant [14]. The total floor space surveyed from 380 food service buildings encompassed 1,819 million square feet in contrast to 157 healthcare facilities comprising a total floor space of 4,155 million square feet [14].

Within hospitals, Singh & Pedamulla (2017) reported that HVAC systems accounted for 44% of energy use in hospitals in India [9], while the ECO-III Project (2009) reported 30-65% of energy within various hospitals are allocated to HVAC function [12]. For other functions, 30-40% of energy usage was for lighting [12]. Barrick et al. (2012) reported 42% of energy in US hospitals are directed to reheat functions [13].

The median building age of healthcare facilities is 29 years [14]. The age of the healthcare facilities can pose an additional challenge to revamping hospital systems and renewal projects that improve energy efficiency because they are both costly and time intensive. Based on 2013 data, India has 19,817 government healthcare facilities and 45,930 private hospitals [9]. The American Hospital Association reported that the US has slightly over 6,000 public and private hospitals [15].

For the purpose of this paper, we focused on inpatient units at acute care hospital facilities. According to the 2012 Commercial Building Energy Consumption Survey (CBECS), the average US inpatient healthcare facility has 23,012 square meters, that operated an average of 168 hours per week [14].

### 2.2 COVID-19 Intervention Measures

According to the Centers for Disease Control and Prevention (CDC), environmental controls are important for decreasing the risk of secondary healthcare associated infections [16]. As potential infection control interventions for COVID-19 are considered, information that pertains to energy consumption at hospitals provides additional context on costs related to implementation of mitigation strategies for person-to-person transmission, as well as environmental contamination. Negative pressured (NP), high efficiency particulate air (HEPA) filtered rooms are defined as areas where air flows directionally towards the inside of a room which prevents airborne and aerosolized droplets from spreading outside the room. Ontario Health Technology Assessment Series (2005) evaluated the cost effectiveness of implementing NP rooms versus combining NP with ultraviolet germicidal irradiation in a single room [17].

In February 2018, Ontario Health Technology Assessment Series’ subsequent report reevaluated the cost impact of using ultraviolet disinfecting devices with mercury versus xenon bulbs [18]. Both the 2005 and 2018 studies evaluated the cost of implementing this type of infrastructure and looked at the budget impact over multiple years. According to the CDC, hospital environmental controls are important for decreasing the risk of secondary (HAI) infections [16]. This paper evaluates the energy implications and impact of NP and XP-UV infection control measures on decreasing secondary transmission of SARS-CoV-2 in acute care hospitals.

### 2.3 Negative Pressure and Portable Air Cleaning Technology as Infection Control Interventions

Doremalen et al. found that viable SARS-CoV-2 virus can remain aerosolized for up to three hours [19]. Rutala et al. (1995) found that portable HEPA (also referred to as portable air cleaners) are able to reduce particles in the air by 90% within a matter of minutes (5-8 minutes) [20]. Control groups, without filtration, took over 171 minutes to reduce air particles [20]. Mead & Johnson (2004) applied portable in-room air cleaners in areas that transformed two to three patient rooms into separate patient rooms, using plastic sheets hung from the ceiling. When using the best configuration, the aerosolized particle counts were 87% lower in the locations of the healthcare workers [21]. Mead & Johnson reported a construction cost of $2,300 ($3,122 in 2020 $USD) per room, that required less than three person-hours to implement [21]. In 2015, Miller et al. demonstrated a hospital’s ability to expediently transform a hospital ward into a negatively pressured surge capacity ward for patients with infections transmitted by aerosolization [22]. They report being able to modify the hospital ward, consisting of 30-beds, in a 40-minute time period. Two HEPA filtered air cleaners were used to expediently create and maintain the negatively pressured space [22].

Loutfy et al. also effectively used 49 expedient, negatively pressured patient rooms to contain the SARS virus in Toronto, Canada [23]. These rooms were modified into negative pressured spaces in a 72-hour time period [23]. Additionally, Loutfy et al. reports similar changes in the Emergency Department, which configured eight, negatively pressured (1,782 square foot) ambulance area within one week [23]. Johnson et al. (2009) reported with an anteroom, these negatively pressured spaces can achieve 99.95% containment, and 99.73% containment during movement throughout these spaces [24]. Garibaldi et al discusses the use of negative pressure and HEPA filters in the implementation of their biocontainment unit reporting HEPA filters within the unit contain 99.99% of particles [25]. Boswell & Fox demonstrated the presence of a portable HEPA filter in the patient room was able to significantly decrease environmental contamination by 90% in room A, 96% in room B, and 75% in room C [26].

### 2.4 Xenon Pulsed Ultraviolet decontamination

Reducing surface decontamination decreases probability of transmission from a viral shedding patient to healthcare worker. Doremalen et al. found that SARS-CoV-2 can remain on surfaces (plastic, stainless, copper, cardboard) ranging from four to 72 hours [19]. Surfaces surrounding infected patients have been found to have the RNA of SARS-CoV-2, even after a 17-day duration [27]. Therefore, the challenge becomes how to effectively decontaminate rooms after discharge of COVID-19 patients. XP-UV (Xenex) uses broad spectrum light (100-280 nm and 380-700 nm) to create bursts of light, which can then sanitize surfaces [28]. Casini found that when pulsed xenon UV light was used in hospital rooms following standard operating cleaning procedures, there was a 100% reduction in pathogens from frequently used surfaces [28].

Simmons et al. reports a 99.97% reduction of SARS-CoV-2 from hard surfaces after exposure to the XP-UV (Xenex) light, after just one minute of light exposure [29]. After two minutes of light exposure, there is greater than 99.997% reduction and at five minutes there is a greater than 99.992% reduction in viruses inoculated on these surfaces [29]. Use of the UV-XP light is also able to reduce inoculated SARS-CoV-2 on N95 respirators by 99.998% (after 5 minutes of light exposure) [29]. Bianco et al. (Italy) used UV-C (200-280 nm) irradiation to evaluate the impact on SARS-CoV-2 viability. After exposure to the UV light, viral replication was inhibited by a factor of 2,000 following 24 hours of light exposure [30]. Dexter et al. also recommends cleaning and treating rooms with UV light to optimize infection control in hospitals, especially as it related to operating rooms [31]. XP-UV has also been demonstrated to statistically reduce the presence of other pathogens in a hospital setting [32, 33].

### 2.5 Alcohol-based sanitizer hand-washing stations

Another risk of transmission is related to the healthcare workers themselves and their ability to infect patients and other healthcare workers [34, 35]. In the Lombardy area of Italy, 20% of healthcare workers became infected [34]. Risk to healthcare workers may be high in the United States as well. With 9,282 COVID-19 patients being healthcare workers, 55% reported the healthcare setting was likely to be their only exposure to COVID-19 [35]. However, most of these healthcare workers did not require hospitalization [35]. Installing hand-washing stations ensures surfaces, and healthcare workers’ hands are clean, which is critical to prevent the transmission of healthcare-associated infections, thus improving the building performance.

The additions of alcohol-based sanitizer dispensers in the corridor (HW for handwashing-stations), was planned to be located adjacent to the door of the treatment room. This convenient location increases the likelihood of healthcare workers (which are assumed to be wearing PPE) sanitizing their hands both before entry and upon exiting an infected patient’s room [36].

Kratzel et al. demonstrated alcohol-based hand rubs are effective against SARS-CoV-2

[37]. Seventy percent ethanol has been found to inactivate live virus in a lab setting

[38]. Location in a convenient, easily visible location increases the use of alcohol-based sanitizer dispensers [36]. Cure & Van Enk found highly visible dispensers, located in the hallways, were the hand-washing stations most frequently used [36].

### 2.6 Intervention-pairs

For this study, the two intervention-pairs evaluated herein are (1) alcohol-based sanitizer dispenser hand-washing stations installed in the hallway, adjacent to patient rooms, combined with either expedient negatively pressured, HEPA filtered patient rooms or (2) xenon-pulsed ultraviolet (XP-UV, Xenex) decontamination cleaning regimens. These intervention-pairs are then evaluated for multiple purposes. The purpose of this paper is to first, evaluate the energy requirements of these infection control intervention-pairs specific to COVID-19, in a hospital setting. The second purpose is to quantify the energy requirements per annum for each intervention, and capture the energy tradeoff for mitigating secondary infections (determine how much energy is needed to prevent x-number of infections). Third, using optimization techniques, assess how capital, operational costs, and the number of secondary infections can be minimized.

## 3. Methods

We first defined the hospital system based on the open-source, Department of Energy hospital data [39]. Three different hospital sizes are evaluated: (1) small hospitals defined as 1-350 patient beds, (2) medium as 351-500, and (3) large hospitals with 501 - 1,000 patient beds. Next, we defined the area of the intervention for small, medium, and large hospitals.

The areas for each intervention to be applied were defined. It is assumed that resident facilities staff will be responsible for procuring materials and augmenting already existing patient rooms with negative pressure and HEPA filtration systems. The NP areas involve emergency room trauma, emergency room exam, operating rooms, intensive care units, and inpatient rooms on Floor 3 and Floor 4, based on the Department of Energy’s representative hospital energy usage data [39].

The NP intervention area incorporates eleven rooms (small hospital), twenty-two rooms (medium hospital), and forty-four rooms (large hospital). The allocation of specific patient areas per hospital size is listed in Table 1. The size (room volume) of these respective patient treatment areas is based on the US Department of Energy hospital data [39]. Twelve air changes per hour (ACH) are assumed to clean 113 cubic meters per hour.

**Table 1.**
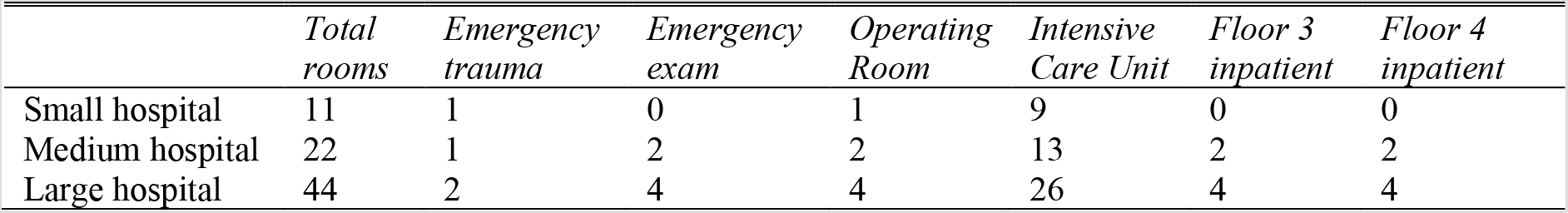
Configuration of expedient Negative pressure, HEPA filtered, patient treatment areas.

The xenon pulsed ultraviolet (XP-UV) decontamination is assumed to incorporate a 15-minute run time per hospital room and five-minute time to relocate to the next location. It is also assumed that existing hospital staff can be trained to operate the XP-UV equipment and that each hospital size will purchase and use one XP-UV robot. The eligible patient areas for decontamination by hospital size are as follows: 76 rooms (small hospital), 143 rooms (medium hospital), and 285 rooms (large hospital). The XP-UV decontamination occurs following standard operating cleaning. Specific hours of usage time by hospital size are listed in Table 2. For the intervention-pairs (NP and HW; XP-UV and HW), we assumed that the number of alcohol-based sanitizer dispensers that are installed equal the number of rooms at that hospital for the NP interventions (11 dispensers in small hospital, 22 dispensers in medium, 44 dispensers in large hospital).

**Table 2.** Number of eligible rooms for decontamination with XP-UV treatment by hospital and hours of usage. It is assumed the UV equipment cleans 113 m^3^ per hour.

|  | *Rooms using XP-UV decontamination* | *Total rooms in hospital* | *Hours used per day* | *No. of XP-UV machines per facility* |
| --- | --- | --- | --- | --- |
| Small hospital | 76 | 253 | 8 | 1 |
| Medium hospital | 143 | 417 | 9 | 1 |
| Large hospital | 285 | 951 | 10 | 1 |

The hospital energy model used in the study to quantify energy usage combines concepts used in Yu et al. (2011), Menezes et al (2014), and Gul et al.(2015) into one integrated model, applied in a healthcare setting [40-42]. Projections for COVID-19 patients were determined using an agent-based, epidemiological model (ABM), which was internally developed to inform bed utilization at Johns Hopkins Medical Institutions healthcare-system and the state of Maryland [43]. The projections from the ABM allowed us to make predictions for the excess influx of COVID-19 patients and inform our energy and cost analysis for pandemic responses in acute care hospitals.

To effectively deliver healthcare services to the community the hospital requires synchronization of various functions. Healthcare delivery is a result of: humans resources management (HRM), financial management (FM), logistics, analysis and imaging, laboratories, patient services, customer relationship management (CRM), risk and compliance, safety, health and environment (SHE), marketing, maintenance, and information and communication technologies (ICT). From an energy perspective, hospital buildings encompass large areas, with substantial heating, ventilation, and air conditioning demands (HVAC), and a plethora of lighting requirements. Hence, to quantify the energy demand of a hospital, all operational requirements and activities must be considered (which will be discussed in further detail).

The modeled hospitals are comprised of the following patient bed counts: small (253 total patient-beds), medium (477 patient-beds), and large hospital (951 patient-beds). Occupancy for normal operations is assumed to be 70% where 10% of occupancy are comprised of walk-ins; occupancy for COVID-19 operations is assumed to be 75% with 15% walk-ins. Three phases of operation during the COVID-19 pandemic operation are modeled which corresponds to the different stages of the pandemic progression. The first phase is during the early stage in which community mitigation strategies are minimal with limited community social distancing. The second phase is a mixed population stage, after the virus has occurred in an area for a longer duration, and community mitigation strategies are beginning to be implemented. The third phase is a late stage which is characterized by declining daily infection counts in a region but new confirmed cases and new hospitalizations are still occurring. The number of applicable rooms where the intervention is being applied varies slightly depending upon the stage of the pandemic in a region (see Tables 4 and 5).

Next five ordinary differential equations from Chamchod & Ruan (2012) were adapted for COVID-19 specific parameters to predict baseline transmission without the intervention(s) [42, 45]. The measure of impact of the transmission is defined by negative pressure, HEPA filter rooms achieving 99.73% containment [24] and the UV-XP achieving a greater than 99.997% reduction in pathogen after two minutes [29].

The progression rate from viral shedding to infection by phase, is based on literature studies reporting COVID-19 transmission rates within a fixed structure. McMichael et al. (2020) provides the progression rate from viral shedding to infection for the early phase [46]; Mizumoto (2020) provides this progression rate for the mixed phase [47]. A combination of a Chamchod & Ruan (2012) equation with Mizumoto infection rate data provides the progression rate for the late phase [44,47]. Capital costs and non-energy related operational costs for each intervention-pair are defined with a cost per respective time frame.

The hospital energy model incorporates all clinical and business processes throughout a hospital (finance, human resources, patient administration, healthcare and emergency delivery, maintenance, patient services, lighting and HVAC operations). The associated energy demands of each of these requirements is incorporated and is related to two databases. The first database involves technical details related to operations involved with all building maintenance activities. The second database includes technical details involving hospital resources (such as personnel, IT, MRIs, and other operational parameters) and the specifics relating to the management of these resources. These databases are then incorporated with the functionality of the floor plans of a hospital. Each patient care area (such as operating rooms, emergency areas, intensive care units, pediatrics, etc.) have specific HVAC and lighting requirements. Therefore, the HVAC and lighting requirements are quantified based on the type of care being provided in that area and associated square meterage.

The hospital energy model is defined for each intervention-pair to determine the energy demand and cost both pre- and post-intervention-pair per year. Subsequently, a mathematical program was applied to determine the optimal investment for each intervention pairs such that infections are minimized. The cost objective function incorporates energy annum costs, capital costs, and non-energy operational and maintenance costs. The output of this objective function is the quantity of pre- and post-intervention infections as well as the optimized percent allocation of funds across the intervention-pairs [45].

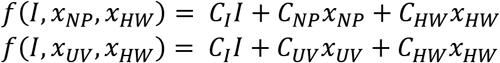

*C_I_ = cost of infection*
*C_NP_ = cost of intervention NP (capital, maintenance, and energy costs)*
*I = number of infections*
*x_NP_ = decision variable for NP intervention, percent allocation to NP*
*C_UV_ = cost of intervention XP-UV (capital, maintenance, and energy costs)*
*x_UV_ = decision variable for UV intervention, % of budget allocation to UV*
*C_HW_ = cost of intervention HW (capital, supply and maintenance costs)*
*X_HW_ = decision variable for HW intervention, % of budget allocation to HW*

**Figure 1.**
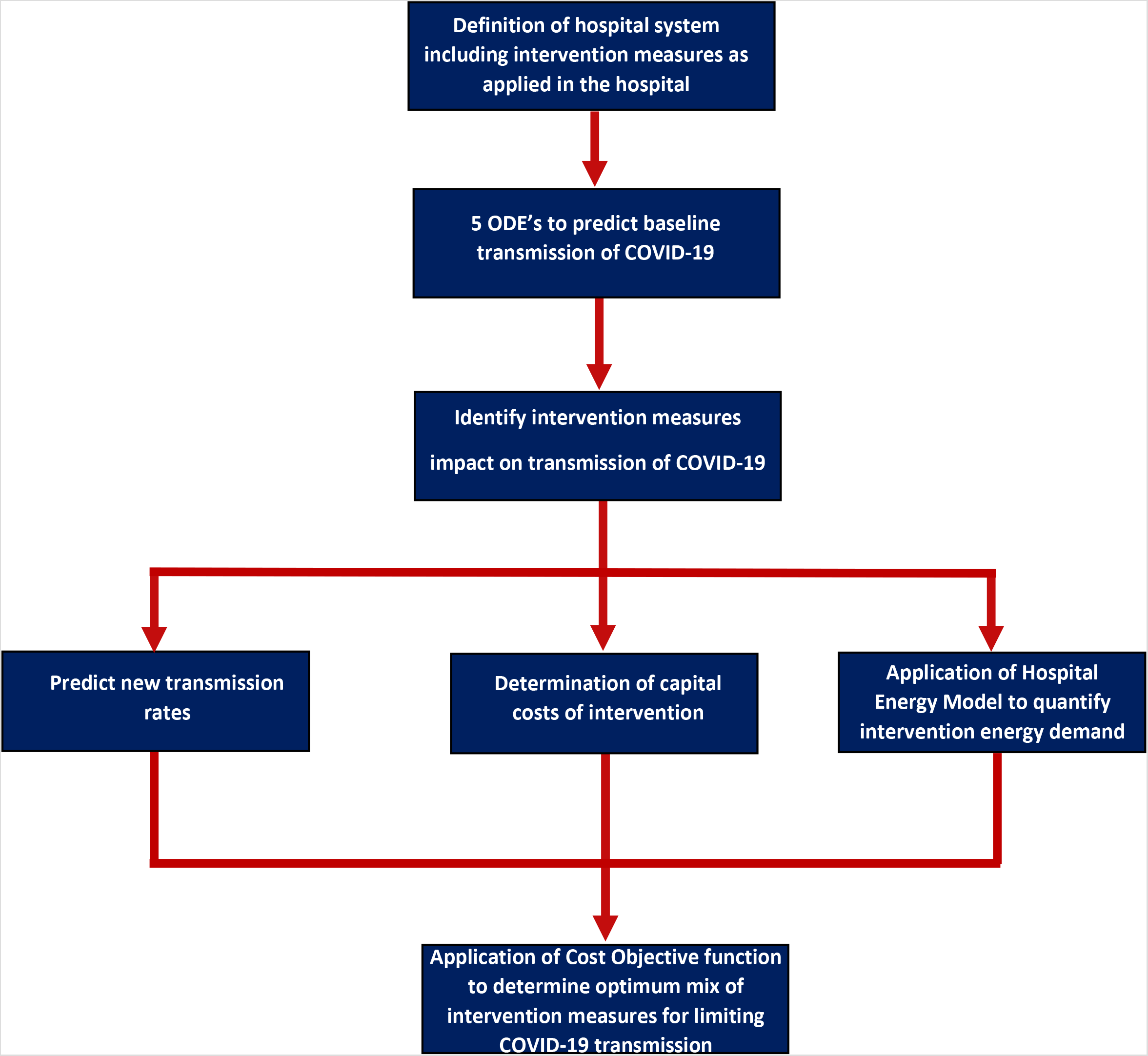
Description of interaction between the interventions, hospitalization, energy, and infection projections.

The objective function minimizes both the cost spent on the interventions and the number of projected secondary infections (I) within the hospital.

## 4. Results

By implementing these interventions early into a pandemic, overall cost-savings are realized by each hospital in this study regardless of size or additional power demand. The cost-saving measures are the infection control interventions which ultimately decreases both financial and infection strain on the hospital. Table 3 outlines the annual energy demand for these hospitals (without either the NP or XP-UV interventions), during both normal and COVID-19 operations. These energy demands are separated by requirement for patient service (which incorporates all clinical activities), IT network, HVAC, and lighting.

**Table 3.** Energy use throughout hospitals without interventions applied

| *Scenario* |  | *Patient Service (kWh)* | *Network (kWh)* | *HVAC (kWh)* | *Lighting (kWh)* |
| --- | --- | --- | --- | --- | --- |
| Normal |  | 587,769 | 318,971 | 1,858,495 | 1,158,696 |
| COVID | Early | 629,246 |  |  |  |
|  | Mixed | 622,953 | 318,971 | 1,858,495 | 1,158,696 |
|  | Late | 613,515 |  |  |  |
| Normal |  | 1,097,361 | 950,576 | 2,688,900 | 1,712,631 |
| COVID | Early | 1,129,014 |  |  |  |
|  | Mixed | 1,106,434 | 950,576 | 2,688,900 | 1,712,631 |
|  | Late | 1,100,789 |  |  |  |
| Normal |  | 2,168,955 | 1,069,145 | 4,948,994 | 3,029,387 |
| COVID | Early | 2,355,822 |  |  |  |
|  | Mixed | 2,238,030 | 1,069,145 | 4,948,994 | 3,029,387 |
|  | Late | 2,190,914 |  |  |  |

**Table 4.** Outputs from Energy analysis and pre- and post-intervention secondary infection quantities, augment select patient rooms with Negative-Pressure

| *Hospital Size* | *Scenario* | *NP Energy Demand (kWh)* | *Total Annual Demand (kWh)* | *% of Annum Energy Demand* | *% of Patient Rooms (intervention/total)* | *Energy Costs* |  |  | *Infections* |  |  | *Cost Savings Per month* |
| --- | --- | --- | --- | --- | --- | --- | --- | --- | --- | --- | --- | --- |
|  |  |  |  |  |  | *Per Annum* | *Additional Occupancy* | *Per Patient Room* | *Without NP* | *With NP* | *Delta* |  |
| Small | Normal | 0 | 3,923,933 | NA | NA | 0 | 0 | Baseline | NA | NA | NA | [39] |
| Small | Covid-Early | 75,219 | 4,040,629 | 1.9% | 4.3%<br>(11/253) | \$7,635 | \$11,845 | \$1,077 | 8 | 2 | 6 | \$229,326 |
| Small | Covid-Mixed | 68,167 | 4,027,285 | 1.7% | 3.6%,<br>9/253, | \$6,919 | \$10,490 | \$1,165 | 7 | 1 | 6 | \$229,326 |
| Small | Covid-Late | 55,239 | 4,004,918 | 1.38% | 2.8%,<br>7/253, | \$5,607 | \$8,220 | \$1,174 | 3 | 1 | 2 | \$76,442 |
| Medium | Normal | 0 | 6,449,470 | NA | NA | 0 | 0 | Baseline | NA | NA | NA | [39] |
| Medium | Covid-Early | 300,877 | 6,782,493 | 4% | 4.6%,<br>22/477 | \$30,539 | \$33,752 | \$1,534 | 17 | 4 | 13 | \$496,873 |
| Medium | Covid-Mixed | 244,462 | 6,703,498 | 4% | 2.7%,<br>13/477 | \$24,813 | \$25,734 | \$1,980 | 16 | 3 | 13 | \$496,873 |
| Medium | Covid-Late | 55,239 | 6,559,128 | 1% | 1.7%,<br>8/477 | \$14,792 | \$15,140 | \$1,682 | 5 | 1 | 4 | \$152,884 |
| Large | Normal | 0 | 11,216,483 | NA | NA | 0 | 0 | Baseline | NA | NA | NA | [39] |
| Large | Covid-Early | 608,805 | 12,012,703 | 5% | 4.6%,<br>44/951 | \$61,794 | \$80,761 | \$1,836 | 30 | 7 | 23 | \$879,083 |
| Large | Covid-Mixed | 484,224 | 11,770,330 | 4% | 2.4%,<br>23/951 | \$49,149 | \$56,160 | \$2,442 | 29 | 7 | 22 | \$840,862 |
| Large | Covid-Late | 238,586 | 11,477,576 | 2% | 1.3%,<br>12/951 | \$24,216 | \$26,445 | \$2,204 | 9 | 1 | 8 | \$305,768 |

**Table 5.** Outputs from Energy analysis and pre- and post-intervention secondary infection quantities, XP-UV.

| *Hospital Size* | *Scenario* | *XP-UV Energy Demand (kWh)* | *Total Annual Demand (kWh)* | *% of Annum Energy Demand* | *% of Patient Rooms (intervention/total)* | *Energy Costs* |  | *Infections* |  |  | *Cost Savings Per Month* |
| --- | --- | --- | --- | --- | --- | --- | --- | --- | --- | --- | --- |
|  |  |  |  |  |  | *Per Annum* | *Per 76 decontam.* | *Without XP-UV.* | *With XP-UV.* | *Inf. Change* |  |
| Small | Normal | 0 | 3,923,933 | NA | NA | 0 | Baseline | NA | NA | NA | [39] |
| Small | Covid-Early | 438 | 3,965,848 | 0.011% | 30%<br>76/253, 24 rm/day | \$45 | \$0.59 | 8 | 1 | 7 | \$267,547 |
| Small | Covid-Mixed | 438 | 3,959,555 | 0.011% | 30%<br>76/253, 24 rm/day | \$45 | \$0.59 | 7 | 1 | 6 | \$229,326 |
| Small | Covid-Late | 438 | 3,950,117 | 0.011% | 30%<br>76/253, 24 rm/day | \$45 | \$0.59 | 3 | 1 | 2 | \$76,442 |
| Medium | Normal | 0 | 6,449,470 | NA | NA | 0 | Baseline | NA | NA | NA | [39] |
| Medium | Covid-Early | 493 | 6,481,616 | 0.0076% | 30%<br>143/477, 27 rm/day | \$50 | \$0.65 | 17 | 3 | 14 | \$535,094 |
| Medium | Covid-Mixed | 493 | 6,459,036 | 0.0076% | 30%<br>143/477, 27 rm/day | \$50 | \$0.65 | 16 | 3 | 13 | \$496,873 |
| Medium | Covid-Late | 493 | 6,453,391 | 0.0076% | 30%<br>143/477, 27 rm/day | \$50 | \$0.65 | 5 | 1 | 4 | \$152,884 |
| Large | Normal | 0 | 11,216,483 | NA | NA | 0 | Baseline | NA | NA | NA | (39) |
| Large | Covid-Early | 548 | 11,403,898 | 0.0048% | 30%<br>285/951, 30 rm/day | \$56 | \$0.73 | 30 | 7 | 23 | \$879,083 |
| Large | Covid-Mixed | 548 | 11,286,106 | 0.0049% | 30%<br>285/951, 30 rm/day | \$56 | \$0.73 | 29 | 5 | 24 | \$917,304 |
| Large | Covid-Late | 548 | 11,283,990 | 0.0049% | 30%<br>285/951, 30 rm/day | \$56 | \$0.73 | 9 | 1 | 8 | \$305,768 |

Energy cost, \$0.10105 per kWh. [48]. Cost per infection \$38,221 [49].

Total demand for these hospitals in normal operations (baseline demand, per annum) is 3,923,932.65 kWh (small), 6,449,470 kWh (medium), and 11,216,483 kWh (large) (Table 3). The cost per kilowatt-hour was $0.1015, based on the commercial energy cost (average cost per kWh of May 2019 and May 2020) in the state of Maryland [48].

Table 4 lists the additional energy usage for the NP intervention (for small, medium, and large hospitals) as: kilowatt-hour(kWh), percentage of total annum hospital energy used for intervention, and additional energy cost per treatment room. The number of secondary infections prevented following the NP intervention (per month) was 6 in a small hospital, 13 in a medium facility, and 22 in a large facility (see Table 3). Using a cost of (hospitalized patient) infection $38,221 per infection [49], the cost savings is determined by multiplying the cost per infection by the (monthly) number of secondary infections prevented. Based on the 2005 Deficit Reduction Act and the Center for Medicare and Medicaid Services policy changes that were effective beginning in 2009, it is assumed that secondary infections (healthcare-associated infections) would not be reimbursed by insurance, Medicare, or Medicaid [50]. Therefore, hospitals and patients would be required to pay for the costs of these treatments. Cost savings are listed in Tables 4 and 5. Varied costs to treat COVID-19 infections have also been reported by Bartsch et al. as $18,579 for hospitalized patients and $3,045 for non-hospitalized patients [51]. It is possible that the cost savings may vary slightly depending on the care given and type of patient.

As depicted in Table 5, the additional annum energy usage cost for the XP-UV (when using 8, 9, and 10 hours a day) intervention in a small, medium, and large hospital is $45, $50, and $56 respectively. The maximum number of infections prevented post-intervention was 6 in a small hospital (per month), 14 in a medium facility (per month), and 24 in a large facility (per month) (see Table 5)

If both intervention-pairs (within this scope of operations), were implemented concurrently, the additional annum energy cost ranges from $8,264 - $11,890 (small hospital), $15,190 - $33,802 (medium), and $26,501 - $80,817 (large hospital).

## 5. Discussion

This study provides a framework which evaluates the energy use of multiple size hospitals during normal and pandemic operations, both with and without infection control interventions. Additionally, we use an Infection De-escalation model to quantify the pre- and post-intervention infection numbers. Optimization techniques are then used to factor in energy use costs, minimize capital costs, and secondary infections within the hospital(s). This study suggests that these flexible, adaptable interventions can be used within hospitals during infection outbreaks to mitigate healthcare-associated infections. The adaptable approach also allows hospital administration and facility staff to adjust the location of their infectious disease surge capacity as needed. These results suggest that these interventions, within the specified scope, can be implemented without large additional energy costs, in a reasonable time frame.

A limitation of the study is that we assumed a 75% occupancy and 15% walk-in percentage during COVID-19 hospital operations. Due to decreases in elective procedures, or low COVID-19 patient admission, hospitals in various areas may not have maintained that occupancy rate. However, the intent was to assess energy usage in locations where hospitals were accommodating maximum patient loads and the associated energy impact and impact of infection control measures. Due to proprietary restrictions, another limitation is that the UV equipment usage was accounted for from a different company than the XP-UV robot itself.

## 6. Conclusions

This study seamlessly combines projections of COVID-19 hospitalizations, evaluation of energy use, and the impact of infection control measures among three different hospital sizes. These frameworks of these systems as well as the measures employed are highly flexible. The expedient NP intervention and XP-UV decontamination equipment can easily be relocated to a different area as needed. This study evaluates a highly adaptable approach to increasing infectious disease surge capacity which could be applied to other diseases as well, such as the H1N1 strain of influenza. Cost savings can be realized in six to eight weeks following intervention implementation. This study suggests that surge capacity can be achieved with a relatively low increase in energy demand and cost.

## Data Availability

Pertinent data is included in the manuscript.

## Acknowledgements

The authors would like to acknowledge Professor Eili Klein, Department of Emergency Medicine, Johns Hopkins University, for his contribution to the community hospitalization epidemiological modeling.

## Appendix Appendix, Cost Listings for Negative Pressure, Xenon-Ultraviolet, and Alcohol-based Sanitizer Stations

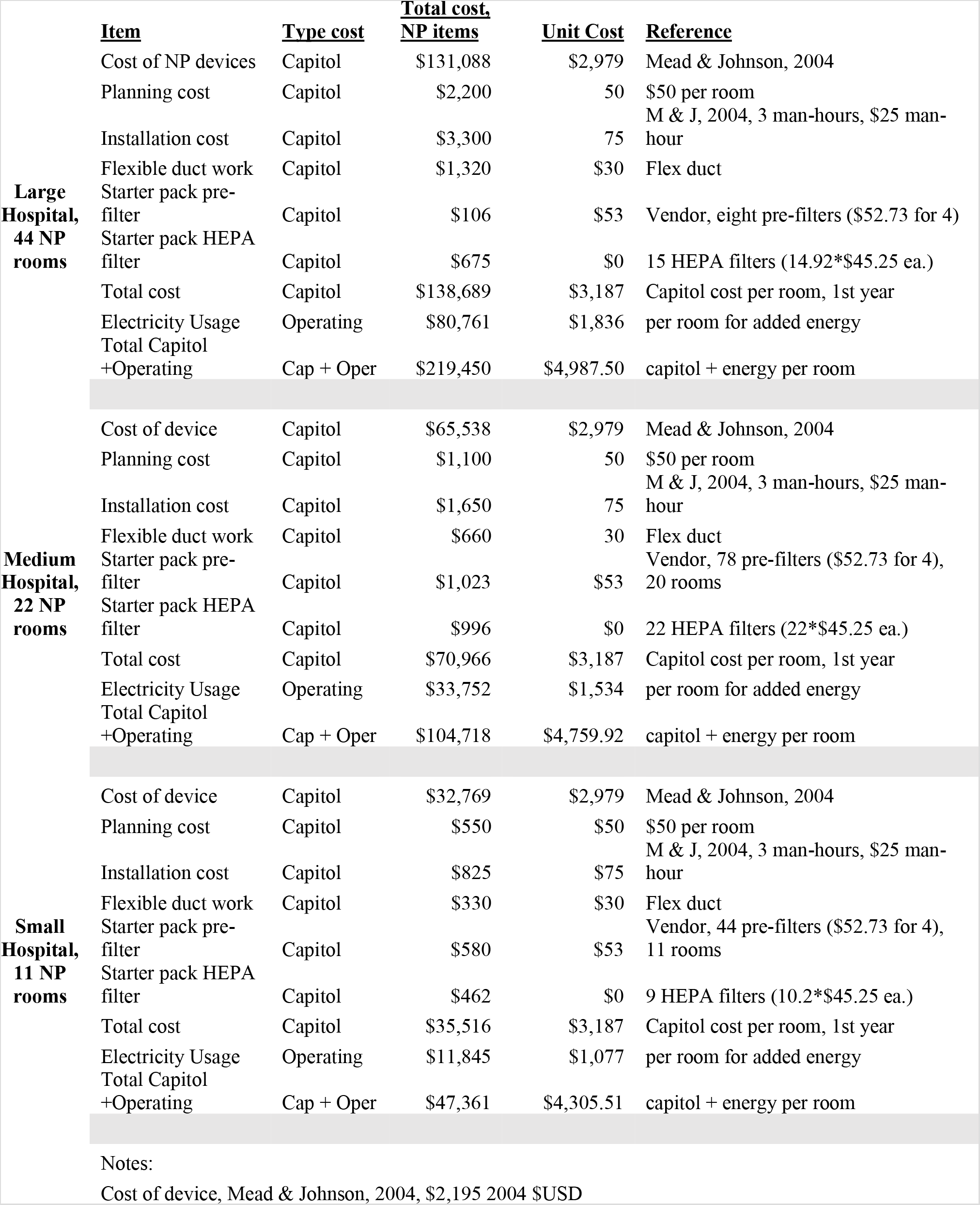
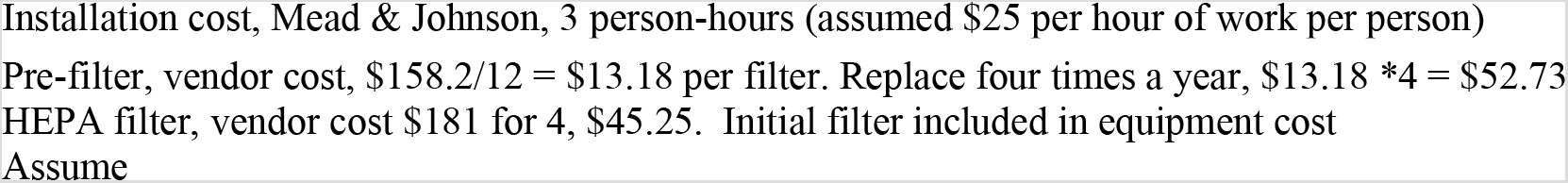

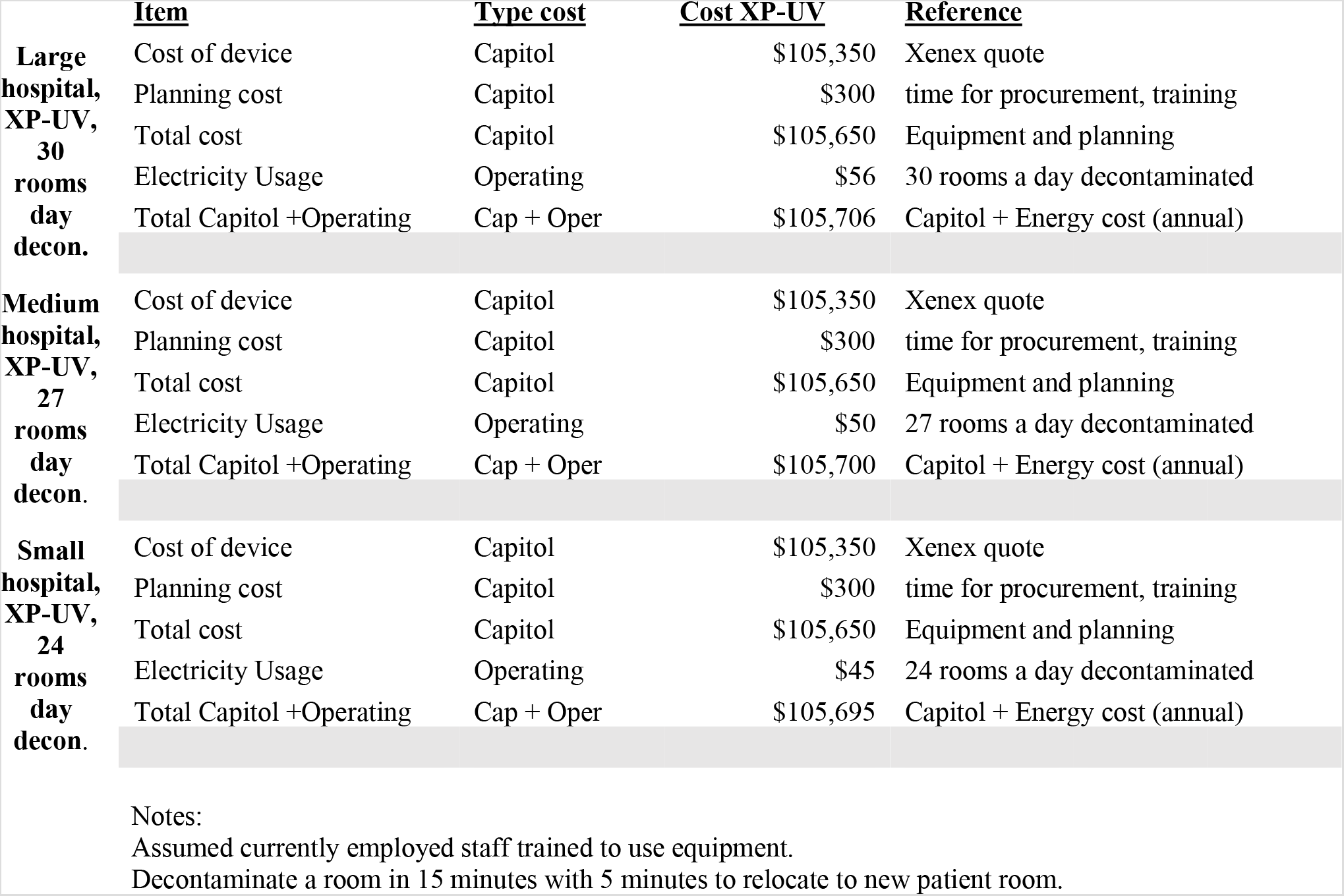

|  | <u>Item</u> | <u>Type cost</u> | <u>Cost XP-UV</u> | <u>Reference</u> |
| --- | --- | --- | --- | --- |
| **Large hospital, XP-UV, 30 rooms day decon.** | Cost of device | Capitol | \$105,350 | Xenex quote |
| | Planning cost | Capitol | \$300 | time for procurement, training |
| | Total cost | Capitol | \$105,650 | Equipment and planning |
| | Electricity Usage | Operating | \$56 | 30 rooms a day decontaminated |
| | Total Capitol +Operating | Cap + Oper | \$105,706 | Capitol + Energy cost (annual) |
| **Medium hospital, XP-UV, 27 rooms day decon.** | Cost of device | Capitol | \$105,350 | Xenex quote |
| | Planning cost | Capitol | \$300 | time for procurement, training |
| | Total cost | Capitol | \$105,650 | Equipment and planning |
| | Electricity Usage | Operating | \$50 | 27 rooms a day decontaminated |
| | Total Capitol +Operating | Cap + Oper | \$105,700 | Capitol + Energy cost (annual) |
| **Small hospital, XP-UV, 24 rooms day decon.** | Cost of device | Capitol | \$105,350 | Xenex quote |
| | Planning cost | Capitol | \$300 | time for procurement, training |
| | Total cost | Capitol | \$105,650 | Equipment and planning |
| | Electricity Usage | Operating | \$45 | 24 rooms a day decontaminated |
| | Total Capitol +Operating | Cap + Oper | \$105,695 | Capitol + Energy cost (annual) |

Notes:
Assumed currently employed staff trained to use equipment.
Decontaminate a room in 15 minutes with 5 minutes to relocate to new patient room.

Assumed currently employed staff trained to use equipment. Decontaminate a room in 15 minutes with 5 minutes to relocate to new patient room.
|  | <u>Item</u> | <u>Type Cost</u> | <u>Total cost</u> | <u>Reference</u> |
| --- | --- | --- | --- | --- |
| **Large Hospital** | Alcohol-based sanitizer dispensers and supplies | Capitol | \$26,044 | Generic Hand Sanitizer Brand Vendor |
| **Medium hospital** | Alcohol-based sanitizer dispensers and supplies | Capitol | \$11,167 | Generic Hand Sanitizer Brand Vendor |
| **Small hospital** | Alcohol-based sanitizer dispensers and supplies | Capitol | \$1,561 | Generic Hand Sanitizer Brand Vendor |

Notes:
Dispensers placed in hallway adjacent to 44 (large hospital), 22 (medium hospital), and 11 (small hospital) patient-rooms [45].
Monetary payment for time spent by healthcare workers to wash hands is accounted for in the total price. All costs listed for interventions are in 2020 \$USD.

## References

[1] N. Christiansen, M. Kaltschmitt, F. Dzukowski. Electrical energy consumption and utilization time analysis of hospital departments and large scale medical equipment, Energy and Buildings. 131 (2016) 172–183.

[2] IEA, Global Energy Review (2020). https://www.iea.org/reports/global-energy-review-2020.

[3] V.C. Broto, J. Kirshner. Energy access is needed to maintain health during pandemics, Nature Energy. 5 (2020) 419–421.

[4] E. Dong, H. Du, L. Gardner. An interactive web-based dashboard to track COVID-19 in real time, The Lancet infectious diseases. 20 (2020) 533–534.

[5] World Health Organization, Coronavirus disease 2019 (COVID-19): situation report, 85 (2020).

[6] C. Coussens, H. Frumkin, Green Healthcare Institutions: Health, Environment, and Economics: Workshop Summary, National Academies Press, 2007.

[7] A. Shajahan, C.H. Culp, B. Williamson, Effects of indoor environmental parameters related to building heating, ventilation, and air conditioning systems on patients’ medical outcomes: A review of scientific research on hospital buildings, Indoor Air. 29 (2019) 161–176.

[8] R.S. Ulrich, C. Zimring, X. Zhu, J. DuBose, H. Seo, Y. Choi, X. Quan, A. Joseph, A review of the research literature on evidence-based healthcare design, HERD: Health Environments Research & Design Journal. 1 (2008) 61–125.

[9] V.K. Singh, L.R.T. Pedamallu, A.C. Ziebell, Addressing Implementation Methods and Challenges for Energy Efficiency in Health Sectors of India, 3rd International Conference on Energy and Environment: bringing together Engineering and Economics Porto, Portugal 29–30 June, 2017 (2017).

[10] A.G. González, J. García-Sanz-Calcedo, D.R. Salgado, A quantitative analysis of final energy consumption in hospitals in Spain, Sustainable Cities and Society. 36 (2018) 169–175.

[11] L. Pérez-Lombard, J. Ortiz, C. Pout, A review on buildings energy consumption information, Energy and Building. 40 (2007) 394–398.

[12] S. Kumar, R. Kapoor, Energy Efficiency in Hospitals: Best Practice Guide, USAID, ECO-III, BEE, India (2009).

[13] J.R. Barrick, R.G. Holdaway, Mechanical Systems: Handbook for Healthcare Facilities, ASHE, 2014.

[14] Energy Information Administration, Commercial buildings energy consumption survey (CBECS) (2015). https://www.eia.gov/consumption/commercial/data/2012/bc/pdf/b1-b2.pdf

[15] American Hospital Association. Fast Facts on Hospitals, AHA Hospital Statistics, 2020 Edition. 2020. https://www.aha.org/system/files/media/file/2020/01/2020-aha-hospital-fast-facts-new-Jan-2020.pdf

[16] L. Sehulster, R.Y. Chinn, Guidelines for environmental infection control in health-care facilities. Recommendations of CDC and the Healthcare Infection Control Practices Advisory Committee (HICPAC)., MMWR. Recommendations and reports: Morbidity and mortality weekly report.Recommendations and reports. 52 (2003) 1–42.

[17] M.A. Secretariat, Air cleaning technologies: an evidence-based analysis, Ontario health technology assessment series. 5 (2005).

[18] Health Quality Ontario, Portable ultraviolet light surface-disinfecting devices for prevention of hospital-acquired infections: a health technology assessment, Ontario health technology assessment series. 18 (2018) 44–48.

[19] N. Van Doremalen, T. Bushmaker, D.H. Morris, M.G. Holbrook, A. Gamble, B.N. Williamson, A. Tamin, J.L. Harcourt, N.J. Thornburg, S.I. Gerber, Aerosol and surface stability of SARS-CoV-2 as compared with SARS-CoV-1, N. Engl. J. Med. 382 (2020) 1564–1567.

[20] W.A. Rutala, S.M. Jones, J.M. Worthmgton, P.C. Reist, D.J. Weber, Efficacy of portable filtration units in reducing aerosolized particles in the size range of Mycobacterium tuberculosis, Infection Control & Hospital Epidemiology. 16 (1995) 391–398.

[21] K. Mead, D.L. Johnson, An evaluation of portable high-efficiency particulate air filtration for expedient patient isolation in epidemic and emergency response, Ann. Emerg. Med. 44 (2004) 635–645.

[22] S.L. Miller, N. Clements, S.A. Elliott, S.S. Subhash, A. Eagan, L.J. Radonovich, Implementing a negative-pressure isolation ward for a surge in airborne infectious patients, Am. J. Infect. Control. 45 (2017) 652–659.

[23] M.R. Loutfy, T. Wallington, T. Rutledge, B. Mederski, K. Rose, S. Kwolek, D. McRitchie, A. Ali, B. Wolff, D. White, Hospital preparedness and SARS, Emerging infectious diseases. 10 (2004) 771.

[24] D.L. Johnson, R.A. Lynch, K.R. Mead, Containment effectiveness of expedient patient isolation units, Am. J. Infect. Control. 37 (2009) 94–100.

[25] B.T. Garibaldi, G.D. Kelen, R.G. Brower, G. Bova, N. Ernst, M. Reimers, R. Langlotz, A. Gimburg, M. Iati, C. Smith, The creation of a biocontainment unit at a tertiary care hospital. The Johns Hopkins medicine experience, Annals of the American Thoracic Society. 13 (2016) 600–608.

[26] T.C. Boswell, P.C. Fox, Reduction in MRSA environmental contamination with a portable HEPA-filtration unit, J. Hosp. Infect. 63 (2006) 47–54.

[27] L.F. Moriarty, Public health responses to COVID-19 outbreaks on cruise ships-worldwide, February-March 2020, MMWR.Morbidity and mortality weekly report. 69 (2020).

[28] B. Casini, B. Tuvo, M.L. Cristina, A.M. Spagnolo, M. Totaro, A. Baggiani, G.P. Privitera, Evaluation of an Ultraviolet C (UVC) Light-Emitting Device for Disinfection of High Touch Surfaces in Hospital Critical Areas, International journal of environmental research and public health. 16 (2019) 3572.

[29] S. Simmons, R. Carrion, K. Alfson, H. Staples, C. Jinadatha, W. Jarvis, P. Sampathkumar, R. Chemaly, F. Khawaja, M. Povroznik, Disinfection effect of pulsed xenon ultraviolet irradiation on SARS-CoV-2 and implications for environmental risk of COVID-19 transmission, medRxiv (2020).

[30] A. Bianco, M. Biasin, G. Pareschi, A. Cavalleri, C. Cavatorta, C. Fenizia, P. Galli, L. Lessio, M. Lualdi, E. Redaelli, UV-C irradiation is highly effective in inactivating and inhibiting SARS-CoV-2 replication, medRxiv (2020).

[31] F. Dexter, M.C. Parra, J.R. Brown, R. W. Loftus, Perioperative COVID-19 Defense: An Evidence-Based Approach for Optimization of Infection Control and Operating Room Management, Infection Control and Operating Room Management. 131 (2020) 37–42.

[32] D.J. Weber, W.A. Rutala, D.J. Anderson, L.F. Chen, E.E. Sickbert-Bennett, J.M. Boyce, Effectiveness of ultraviolet devices and hydrogen peroxide systems for terminal room decontamination: focus on clinical trials, Am. J. Infect. Control. 44 (2016) e77-e84.

[33] M.M. Nerandzic, P. Thota, T. Sankar, A. Jencson, J.L. Cadnum, A.J. Ray, R.A. Salata, R.R. Watkins, C.J. Donskey, Evaluation of a pulsed xenon ultraviolet disinfection system for reduction of healthcare-associated pathogens in hospital rooms, Infection Control & Hospital epidemiology. 36 (2015) 192–197.

[34] A. Remuzzi, G. Remuzzi, COVID-19 and Italy: what next?, The Lancet (2020).

[35] S.L. Burrer, M.A. de Perio, M.M. Hughes, D.T. Kuhar, S.E. Luckhaupt, C.J. McDaniel, R.M. Porter, B. Silk, M.J. Stuckey, M. Walters, Characteristics of health care personnel with COVID-19-United States, February 12-April 9, 2020, MMWR. 69 (2020) 477–481.

[36] L. Cure & R. Van Enk, Effect of hand sanitizer location on hand hygiene compliance, American Journal of Infection Control, 43 (2015) 917–921.

[37] A. Kratzel, D. Todt, P. V’kovski, S. Steiner, M. Gultom, T.T.N. Thao,N. Ebert, M. Holwerda, J. Steinmann, D. Niemeyer, Inactivation of severe acute respiratory syndrome coronavirus 2 by WHO-recommended hand rub formulations and alcohols, Emerging Infect. Dis. 26 (2020).

[38] A.W. Chin, J.T. Chu, M.R. Perera, K.P. Hui, H. Yen, M.C. Chan, M. Peiris, L.L. Poon, Stability of SARS-CoV-2 in different environmental conditions, The Lancet Microbe. 1 (2020) e10.

[39] K.C. Hallett, Office of Energy Efficiency & Renewable Energy (EERE), Commercial Reference Building: Hospital, US Department of Energy. Accessed August 2019 from https://openei.org/wiki/Commercial_Reference_Buildings#Building_Types;https://openei.org/datasets/dataset/commercial-reference-building-hospitalhttps://www.energy.gov/eere/downloads/reference-buildings-building-type-hospital; Database license, Open Data Commons Attribution License 1.0.

[40] Z. Yu, B.C. Fung, F. Haghighat, H. Yoshino, E. Morofsky, A systematic procedure to study the influence of occupant behavior on building energy consumption, Energy Build. 43 (2011) 1409–1417.

[41] A.C. Menezes, A. Cripps, R.A. Buswell, J. Wright, D. Bouchlaghem, Estimating the energy consumption and power demand of small power equipment in office buildings, Energy Build. 75 (2014) 199–209.

[42] M.S. Gul, S. Patidar, Understanding the energy consumption and occupancy of a multi-purpose academic building, Energy Build. 87 (2015) 155–165.

[43] Klein, 2020. Internal Maryland state, Community-Based Hospitalization Model. Johns Hopkins School of Medicine, Department of Emergency Medicine, Internal Medicine. Spring 2020.

[44] F. Chamchod, S. Ruan, Modeling methicillin-resistant Staphylococcus aureus in hospitals: transmission dynamics, antibiotic usage and its history, Theoretical Biology and Medical Modelling. 9 (2012) 25.

[45] M. M. Squire, T. Igusa, S. Siddiqui, G.K. Sessel, E.N. Squire Jr, Cost-Effectiveness of Multifaceted Built Environment Interventions for Reducing Transmission of Pathogenic Bacteria in Healthcare Facilities, Health Env Research & Design Journal, 12 (2019) 147–161.

[46] T.M. McMichael, D.W. Currie, S. Clark, S. Pogosjans, M. Kay, N.G. Schwartz, J. Lewis, A. Baer, V. Kawakami, M.D. Lukoff, Epidemiology of Covid-19 in a long-term care facility in King County, Washington, N. Engl. J. Med. 382 (2020) 2005–2011.

[47] K. Mizumoto, K. Kagaya, A. Zarebski, G. Chowell, Estimating the asymptomatic proportion of coronavirus disease 2019 (COVID-19) cases on board the Diamond Princess cruise ship, Yokohama, Japan, 2020, Eurosurveillance. 25 (2020) 2000180.

[48] EIA, Commercial Energy Cost, Maryland, 2019, 2020. Accessed from https://www.eia.gov/electricity/monthly/epm_table_grapher.php?t=epmt_5_6_a

[49] FAIR Health Report, The Projected Economic Impact of the COVID-19 Pandemic on the US Healthcare System, March 25, 2020,13. Accessed from https://www.fairhealth.org/article/fair-health-releases-brief-on-covid-19

[50] CMS, 2020. https://www.cms.gov/Medicare/Medicare-Fee-for-Service-Payment/HospitalAcqCond/Hospital-Acquired_Conditions

[51] S.M. Bartsch, M.C. Ferguson, J.A. McKinnell, K.J. O’Shea, P.T. Wedlock, S.S. Siegmund, B.Y. Lee, The Potential Health Care Costs And Resource Use Associated With COVID-19 In The United States: A simulation estimate of the direct medical costs and health care resource use associated with COVID-19 infections in the United States, Health Aff. (2020) 10.1377/hlthaff.2020.00426.

